# Characteristics of head and neck alignment and function of deep cervical flexor muscles in patients with nonspecific neck pain

**DOI:** 10.1101/2022.12.07.22283100

**Authors:** Tomoko Kawasaki, Shunsuke Ohji, Junya Aizawa, Tomoko Sakai, Kenji Hirohata, Taiichi Koseki, Hironobu Kuruma, Atsushi Okawa, Tetsuya Jinno

## Abstract

**Objective:** To compare forward head posture (FHP) in natural and corrected head postures between patients with nonspecific neck pain (NSNP) and controls, and to clarify the relationship between natural- and corrected-head posture angle differences and deep cervical flexor function.

**Design:** Survey study

**Setting:** Patients with NSNP were outpatients at an orthopedic clinic; the controls were community volunteers.

**Participants:** Thirty-eight patients were divided into the NSNP and control groups (n=19 each), including patients reporting a numerical rating pain score of 3-7 for at least 3 months and those with no neck pain within 12 months previously, respectively.

**Interventions:** To evaluate FHP, the cranial rotation and vertical angles were measured using lateral photographs of the head and neck. The craniocervical flexion test was used to evaluate deep cervical flexor activation and endurance.

**Main outcome measures:** We evaluated the head and neck alignment in natural and corrected head postures and the relationship between the degree of change and deep cervical flexor function.

**Results:** The FHP in the natural head posture did not differ significantly between the groups. For corrected head posture, the FHP was significantly smaller in the NSNP group than in the control group. In the NSNP group, the cranial rotation and vertical angles were significantly different between natural and corrected head postures, and the angle difference between these postures was significantly correlated with deep cervical flexor function.

**Conclusions:** In patients with NSNP, hypercorrection in the corrected head posture was shown and may be correlated with dysfunction of the deep cervical flexors. Further investigation into the causal relationship between hypercorrection, deep neck flexor dysfunction, and neck pain is required.

---

Approximately 70% of the global population will experience nonspecific neck pain (NSNP) at some time in their lives.^1^ NSNP is a social issue as it places a large burden on healthcare systems and results in a loss of productivity.^2,3^ To improve this, it is necessary to clarify the factors related to NSNP.

One factor known to be related to NSNP is head and neck malalignment in the sagittal plane, particularly forward head posture (FHP). A cross-sectional study that measured head posture in the sagittal plane found a correlation between FHP and neck pain intensity.^4^ In systematic reviews that investigated the relationship between neck pain and FHP, some reported that the occurrence of FHP as the natural head posture was different between patients with neck pain and the control groups.^5^ In contrast, others reported that there was no difference between the two groups.^6^ Even though these studies considered FHP as a relevant factor associated with NSNP, no consensus has been reached.

The characteristics of head and neck alignment among patients with NSNP may not be easily detected by simply analyzing the natural head posture. A systematic review on proprioceptive dysfunction in patients with neck pain found that joint positional error in the neck is related to neck pain.^7^ Another study found that measurement of cervical joint positional proprioception in the horizontal and sagittal planes resulted in significant errors during head repositioning among those with neck pain compared to those without.^8^ These studies suggest that the characteristics of head and neck alignment in patients with NSNP may be better understood by not only evaluating natural head posture but also considering alignment during a task involving corrected head posture. No previous study has measured head and neck alignment in natural and corrected head postures and simultaneously analyzed these characteristics in patients with NSNP.

In clinical settings, hypercorrection is frequently observed in patients with NSNP when instructed to correct their head posture. For this reason, the deep cervical flexors may be dysfunctional in these patients. A study of the relationships between FHP, using x-ray images, and neck muscle function among college students reported a negative correlation between FHP and endurance of the deep cervical flexors.^9^ Patients with NSNP frequently present with dysfunction of the deep cervical flexors,^10,11^ which tends to lead to compensatory activation of the surface muscles of the neck.^10–12^ These studies suggest that in patients with NSNP, the functional impairment of the deep cervical flexors, which contribute to the correction of FHP, may lead to compensatory activity of the surface muscles of the neck, causing hypercorrection. However, the relationship between the corrected head posture and the function of the deep cervical flexors is unclear.

Therefore, this study aimed to compare the extent of FHP in natural and corrected head postures between the NSNP and control groups and clarify the relationship between the angle difference (corrected - natural) and the function of the deep cervical flexors. We hypothesized that: there are no differences in FHP in the natural head posture between the NSNP and control groups, the corrected head position results in less FHP and further change from the natural head posture in the NSNP group compared to the control group, and there is a relationship between the angle difference between natural and corrected head postures and the function of the deep cervical flexors.

## Methods

### Procedures

This was a survey study. Participant demographics and pain intensity scores were collected using a self-descriptive questionnaire. Lateral head and neck photographs were taken, and the deep cervical flexors functions were assessed using the craniocervical flexion test (CCFT). All participants provided written informed consent before initiation of the study. This study was approved by the university’s Institutional Review Board (approval no. M2019-040).

### Participants

We defined the participant groups as follows: the NSNP group and control group. Inclusion criteria for the patients in the NSNP group were those who were examined at an orthopedic clinic due to neck pain and reported a pain score on the numerical rating scale (NRS) of 3-7, for at least 3 months.^13–15^ The control group included individuals who reported no neck pain (NRS 0) in the 12-month period prior to the measurement.^14^ The exclusion criteria were as follows: history of diseases of the spine, rheumatoid arthritis, previous spine surgery, pain symptoms in the neck or shoulder area, and pregnancy. Patients in the NSNP group were recruited from outpatients attending a single orthopedic clinic. Participants in the control group were recruited from volunteers; recruitment was conducted by poster advertisement. The sample size was calculated using G*Power^16^ based on data from similar previous studies,^4,17,18^ with a minimum sample size of 19 in each group for a total of 38 (effect size (d) = 0.96; alpha = 0.05; power = 0.8; two-tailed).

### Evaluation of Pain Intensity

The intensity of neck pain was measured using the NRS.^19^ The average intensity of pain experienced during the week before the study was measured using a self-descriptive questionnaire.^20^ The inter-rater reliabilities of this method were high (intraclass correlation coefficient [ICC] = 0.76).^21^

### Measurement of Head and Neck Angles

To measure FHP, lateral head and neck photographs of the participants were taken. Participants were seated with the head and trunk in the upright position and asked to gaze forward; the height of the chair was 40 cm. The arms were extended, and the hands were placed on either side of the body. The measurement task involved the participant seated with voluntary natural head posture (Figure 1a) followed by corrected head posture (Figure 1b). The corrected head posture was the position where the participant felt that the head was positioned directly above the trunk. Lateral photographs of the head and neck were taken using a digital video camera (HDR-CX720V; Sony Corp., Tokyo, Japan). The distance between the camera lens and the participant was 300 cm.^22,23^ The height of the camera lens was adjusted to the level of the lateral canthus of the participant, and the lateral canthus was captured in the center of the image. Photographs were taken once in the natural head posture and the corrected head posture, respectively.

**Figure 1.**
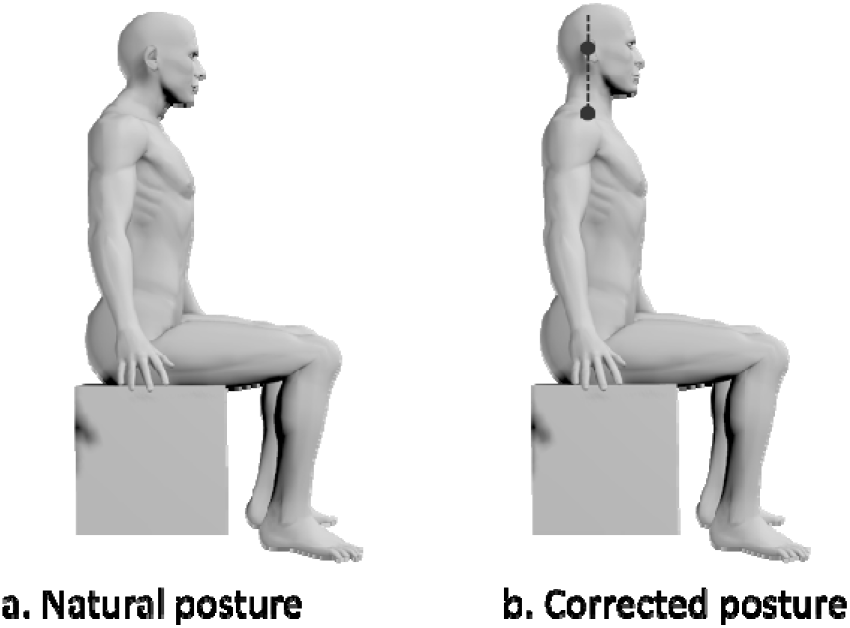
Measurement task During the measurement task, the patient was seated with the head in the voluntary natural and corrected postures.

### Photographic Analysis

To evaluate FHP, the cranial rotation angle (CRA) and cranial vertical angle (CVA) were measured using the lateral photographs (Figure 2).^9^ The CRA was determined by measuring the angle between the line connecting C7 with the tragus of the ear and the line connecting the tragus of the ear with the lateral canthus of the eye.^24^ The CVA was calculated by measuring the angle between the line connecting C7 with the tragus of the ear and the horizontal line.^24^ The intra- and inter-rater reliabilities of these methods were high (ICC = 0.86-0.96).^25–27^ Each photograph was measured by the same physical therapist, who specialized in orthopedic rehabilitation for 12 years, using ImageJ software (National Institutes of Health, Bethesda, Maryland, USA).

**Figure 2.**
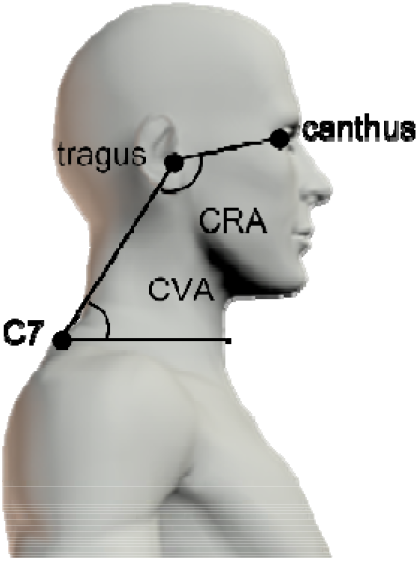
Photographic analysis The head and neck angles (CRA and CVA) were measured to evaluate the forward head posture using lateral photographs. CRA, cranial rotation angle; CVA, cranial vertical angle

### Craniocervical Flexion Test (CCFT)

The CCFT^28, 29^ was used to evaluate the activation and endurance of the deep cervical flexors. In the supine position, a sphygmomanometer cuff was placed behind the participant’s neck in a position where it abutted the occiput. It was then inflated to a stable baseline pressure of 20 mmHg, which is the standard pressure sufficient to fill the space between the testing surface and neck (Figure 3). The movement was performed gently and slowly as a head nodding action (as if saying “yes”) for five incremental increasing stages; 2 mmHg progressive pressure increases from the baseline of 20 mmHg to a maximum of 30 mmHg, with each stage being held for 10 seconds. The CCFT was performed through two phases; in the first phase, we measured the maximum pressure the participant could hold for 10 seconds with upper cervical flexion, and this was used as the activation pressure score. Next, the number of times the participant could repeat the 10 second holding exercise (up to a maximum of 10 times) was recorded, and the activation pressure score was set as the target pressure. The performance pressure index was calculated by multiplying the activation pressure score by the number of successful repetitions as the endurance of the deep cervical flexors. During CCFT, the deep cervical flexors, especially the longus colli and longus capitis muscle, are activated, and the electromyogram amplitude increases with each stage of the test.^30,31^ The inter-rater reliability of this method was high (ICC = 0.81-0.93).^11,29^

**Figure 3.**
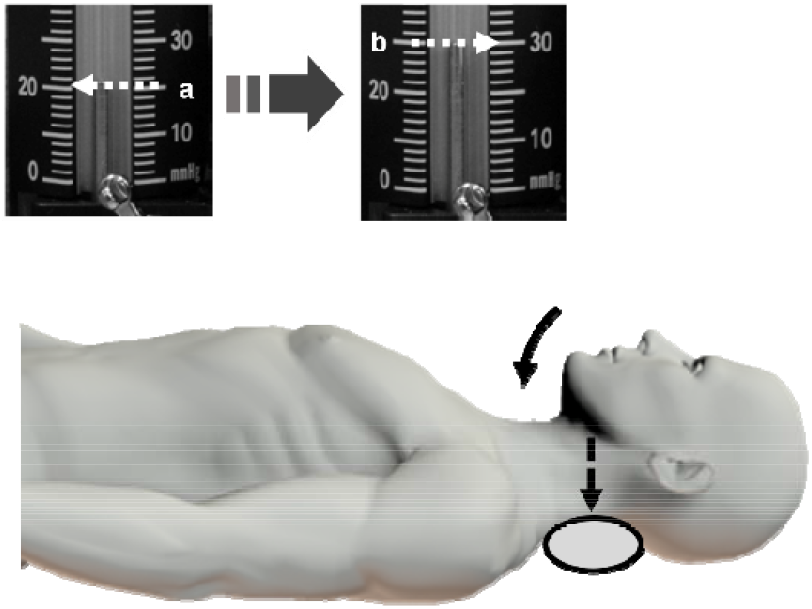
Craniocervical flexion test The craniocervical flexion test was used to evaluate the activation and endurance of the deep cervical flexors. (a) stable baseline pressure of 20 mmHg (b) Five incremental increasing stages (2 mmHg progressive pressure increases from the baseline of 20 mmHg to a maximum of 30 mmHg)

### Calculated Data

The CRA and CVA in the natural and corrected head postures, measured using ImageJ in lateral photographs, were used for the analysis. The activation pressure score and (performance index) PI measured by CCFT were also analyzed. The angle difference between natural and corrected head postures of CRA and CVA (corrected - natural = Difference-CRA, Difference-CVA) was also calculated.

### Statistical Analysis

The normality of each variable was confirmed using the Shapiro–Wilk test. Normally distributed data were summarized as mean and standard deviations and analyzed using the *t*-test or Pearson’s correlation coefficient. Non-normally distributed data were summarized as medians and interquartile ranges and analyzed using the Wilcoxon signed-rank test, Mann–Whitney U test, or Spearman’s rank correlation coefficient. CRA and CVA in natural and corrected head postures and PI were compared between the NSNP and control groups using the unpaired *t*-test. For the comparison of activation pressure scores between the NSNP and control groups, the Mann–Whitney U test was used. Inter-group comparisons of CRA and CVA between natural and corrected head postures were analyzed using the paired *t*-test. The relationship between the difference-CRA or difference-CVA and activation pressure score or PI was analyzed using Spearman’s rank correlation coefficient.

All data were analyzed using SPSS (version 21.0; IBM Corp, Armonk, NY, USA). Statistical significance was set at P<0.05. The effect size (*t*-test = d; Wilcoxon’s signed rank test, Mann–Whitney=*r*) was calculated for each variable.

## Results

A total of 38 participants completed the measurement: nineteen patients with NSNP and nineteen controls were included in the study (Table 1). There were no significant differences in CRA and CVA in the natural head posture between the NSNP and control groups (P = 0.39, d = 0.29; P = 0.86, d = 0.06) (Table 2). The CRA in the corrected head posture was significantly smaller in the NSNP group than in the control group (P < 0.05, d = 1.00) (Table 2). The CVA in the corrected head posture was significantly larger in the NSNP group than in the control group (P < 0.01, d = 1.08) (Table 2). In the NSNP group, there was a significant difference in the CRA and CVA between natural and corrected head postures (P < 0.01, d = 1.05; P < 0.01, d = 1.39) (Table 3). In contrast, there was no significant difference in the control group in CRA and CVA between natural and corrected head postures (P = 0.70, d = 0.05; P = 0.08, d = 0.28) (Table 3). The activation pressure score and PI were significantly lower in the NSNP group than in the control group (P < 0.05 *r* = −0.39; P < 0.01, d = 1.03) (Table 4). In the NSNP group, the difference-CRA and activation pressure score or PI had a significant positive correlation (ρ = 0.60, P< 0.01; ρ = 0.76, P< 0.01), and the difference-CVA and activation pressure score had a significant negative correlation (ρ = −0.54, P< 0.05) (Table 5).

**Table 1.** Participant characteristics.

|  | NSNP group | Control group |
| --- | --- | --- |
| Men:women | 2:17 | 10:9 |
| Age (years) | 48.0 (13.0) | 36.0 (30.0) |
| Height (cm) | 160.8±6.1 | 165.4±9.9 |
| Weight (kg) | 49.0 (7.5) | 63.0 (13.0) |
| Body mass index | 19.3 (1.5) | 22.9 (1.7) |
| Numerical rating scale score | 4.0 (3.0) | 0.0 (0.0) |

Height is reported as mean±SD
Other data are reported as median (interquartile range)

**Table 2.**
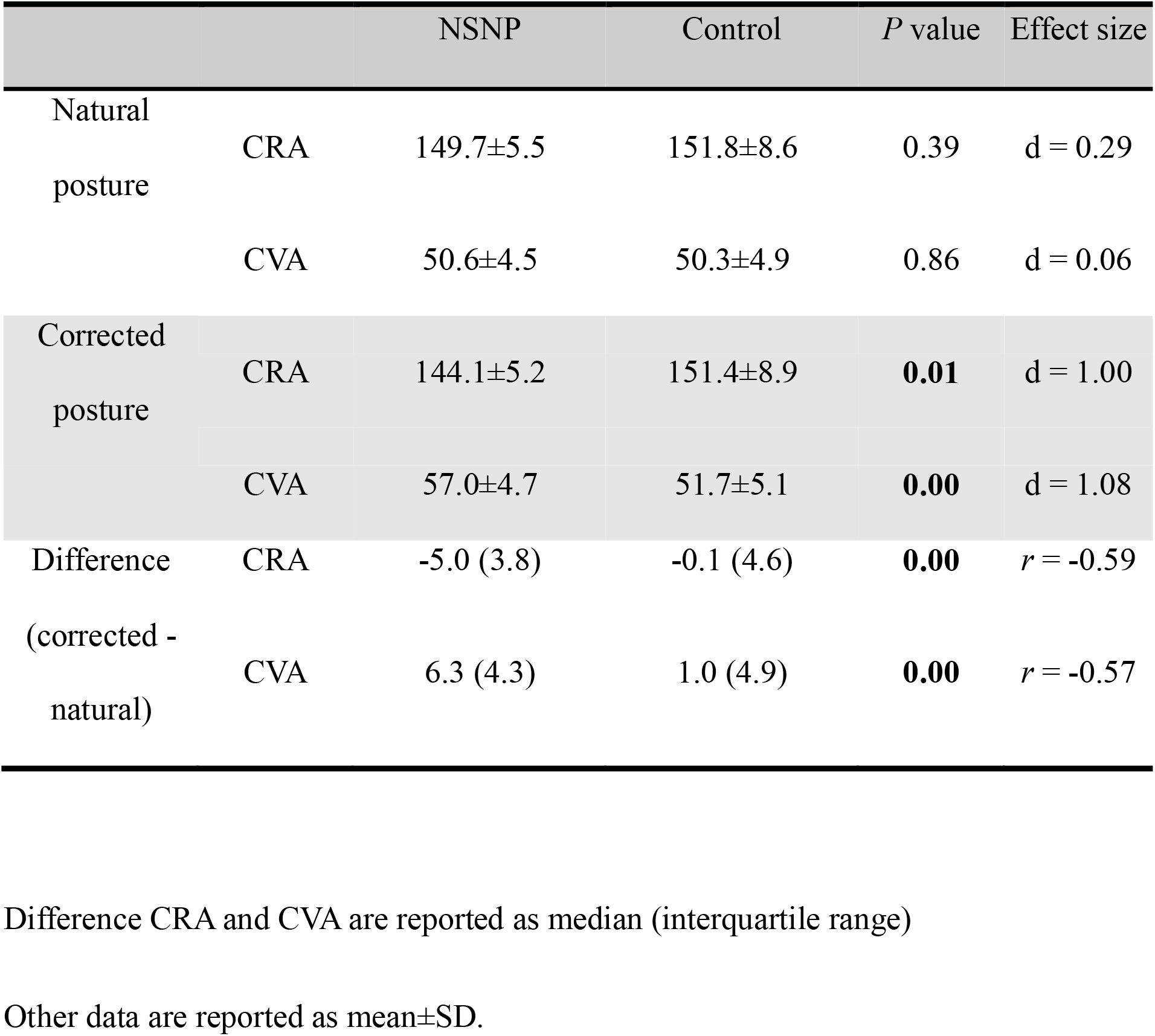
Comparison of head and neck alignment between patients with NSNP and the control group.

**Table 3.**
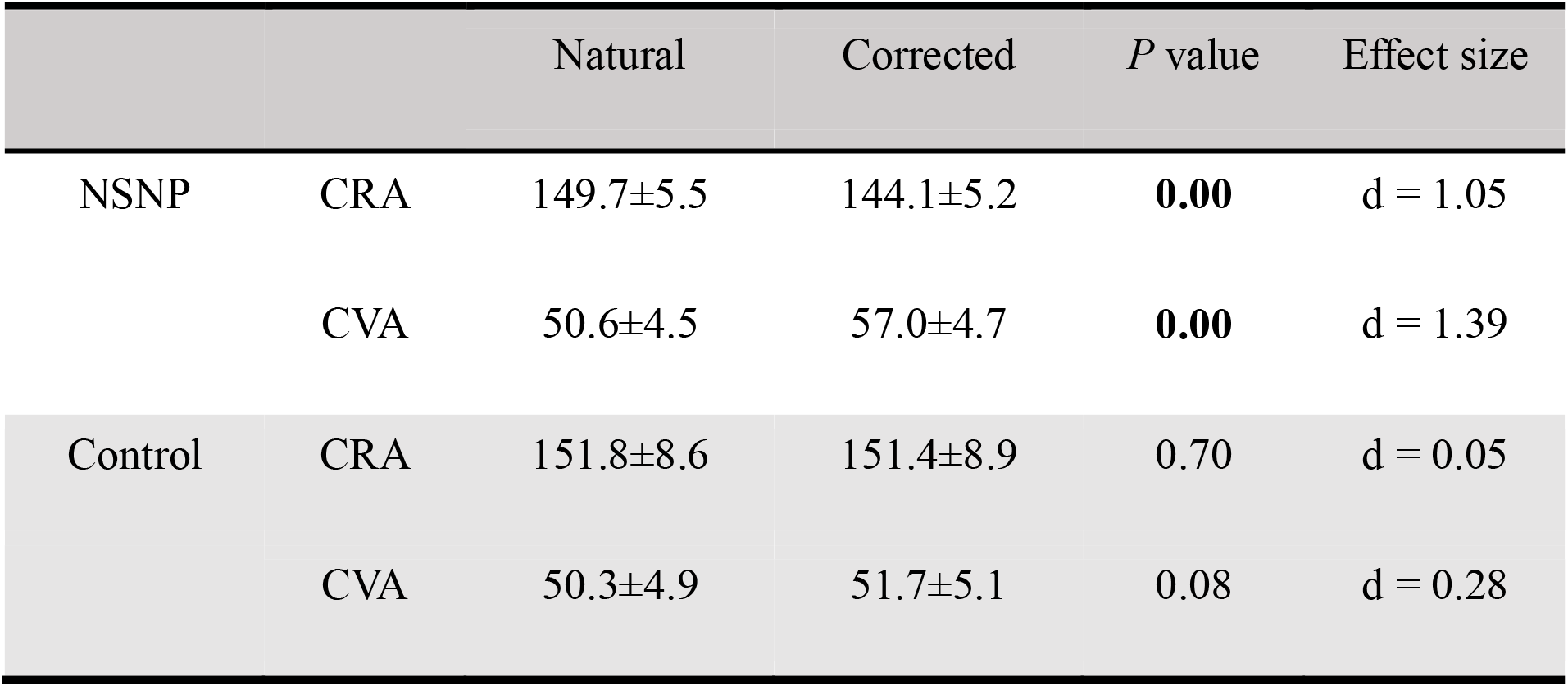
Comparison of head and neck alignment between natural and corrected.

|  |  | Natural | Corrected | *P* value | Effect size |
| --- | --- | --- | --- | --- | --- |
| NSNP | CRA | 149.7±5.5 | 144.1±5.2 | **0.00** | d = 1.05 |
|  | CVA | 50.6±4.5 | 57.0±4.7 | **0.00** | d = 1.39 |
| Control | CRA | 151.8±8.6 | 151.4±8.9 | 0.70 | d = 0.05 |
|  | CVA | 50.3±4.9 | 51.7±5.1 | 0.08 | d = 0.28 |

**Table 4.** Comparison of function of deep cervical flexor muscles in NSNP and control groups.

|  | NSNP | Control | *P* value | Effect size |
| --- | --- | --- | --- | --- |
| Active pressure score | 5.0 (5.0) | 6.0 (3.0) | 0.02 | $r = -0.39$ |
| Performance index | 37.5±23.8 | 61.3±22.2 | 0.00 | $d = 1.03$ |

**Table 5.** Relationship between the degree of change from natural to corrected head postures and the function of deep cervical flexor muscles.

|  | All |  | NSNP |  | Control |  |
| --- | --- | --- | --- | --- | --- | --- |
|  | D-CRA | D-CVA | D-CRA | D-CVA | D-CRA | D-CVA |
| Active pressure score | 0.49* | -0.40<sup>†</sup> | 0.60* | -0.54<sup>†</sup> | 0.25 | 0.24 |
| Performance index | 0.49* | -0.23 | 0.76* | -0.33 | -0.01 | 0.42 |

\*\* <0.01, <sup>†</sup> <0.05
D, Difference (corrected-natural)

## Discussion

We compared CRA and CVA in natural and corrected head postures between patients with NSNP and controls. In the natural head posture, there was no difference in CRA and CVA between the NSNP and control groups. In the NSNP group, the CRA in the corrected head posture was smaller, the CVA was larger, and the angle difference between natural and corrected head postures was bigger than in the control group. We also found relationships between the angle differences between natural and corrected head postures and the function of the deep cervical flexors in the NSNP group. The results of this study support our hypothesis.

In the natural head position, there was no difference in the CRA and CVA between the NSNP and control groups. In the corrected head position, CRA was smaller and CVA larger in the NSNP group than in the control group. Goniometric measurements of the CVA during standing in patients with neck pain and healthy participants showed no differences between the two groups.^18^ In contrast, a study of head and neck alignment with a simulated computer workstation in patients with neck pain and healthy controls, using photographs taken by a digital camera, showed that the CVA of the neck pain group was significantly smaller than that of the healthy controls.^32^ While comparing head and neck alignment in the natural head posture between the NSNP and control groups, previous study results are conflicting regarding the difference between the groups; this may be influenced by the measurement environment and the task performed. No previous studies have compared head and neck alignment in the corrected head postures between patients with NSNP and healthy controls. The results of this study suggest that differences in head and neck alignment between patients with NSNP and healthy controls are characterized in the corrected head posture. Therefore, when evaluating the characteristics of head and neck alignment of patients with NSNP, it is better to observe not only the natural head posture but also corrected head posture.

The head and neck alignment of the NSNP group showed no difference in the natural head posture compared to the control group, but there was a significant difference in the corrected head posture; the angle difference between natural and corrected head postures was larger. A study involving measurement of the head and neck alignment in the natural head posture in healthy participants showed the CRA was 148.9 (8.7)° and the CVA was 50.9 (3.8)°.^33^ A study measuring natural head posture using a digital camera in healthy participants reported the CVA as 50.4 ± 5.2°.^34^ In the corrected head and neck alignment (CRA: 149.7±5.5°, CVA: 50.6±4.5°) of the NSNP group in this study, the CRA was smaller and CVA larger than those in previously reported healthy data. The CRA is reduced by flexion of the upper cervical spine, while the CVA is increased by extension of the middle-lower cervical spine; these combined movements lead to a retracted head posture.^33^ Collectively, these findings suggest that patients with NSNP have a larger degree of flexion of the upper cervical spine and extension of the middle-lower cervical spine in the corrected head position from the natural head posture, showing a hypercorrected head posture compared to control groups.

One factor leading to hypercorrection of head and neck alignment in patients with NSNP may be dysfunction of the deep cervical flexors. This study showed a correlation between the angle difference between natural and corrected head postures and the function of the deep cervical flexors in the NSNP group. Comparing the function of the deep cervical flexors using the CCFT in patients with cervical pain and healthy controls, Juul et al. reported that activation pressure scores and PI were significantly lower in the cervical pain patient group.^10,11^ In patients with NSNP, the surface muscles tend to compensate for the dysfunction of the deep cervical flexors with upper cervical flexion.^12^ The surface muscles have longer moment arms than the deep muscles and are poorly adapted to segmental and micro-movements.^35^ Thus, dysfunction of the deep cervical flexors may lead to hypercorrection in the corrected head posture.

The characteristic head and neck alignment of patients with NSNP may not be accurately detected by evaluating natural head posture only. It is necessary to also observe the corrected head posture when evaluating the head and neck alignment of patients with NSNP in the sitting position. The corrected head posture of patients with NSNP is likely to hypercorrect posteriorly, related to dysfunction of the deep cervical flexors. If hypercorrection of the posture is observed in patients with NSNP, treatment might be aimed at improving the head posture and function of the deep cervical flexors to reduce neck pain. Evaluating the corrected head posture and the function of the deep cervical flexors may be valuable in instructing patients with NSNP to attempt and correct their head posture, which might ultimately aid in reducing their pain.

### Study Limitations

This study has several limitations. Because of the unequal ratios of male and female participants in both groups, the effect of sex is not known. Women are at higher risk for NSNP^36^ and have larger FHP than men,^23^ but the causal relationship is unknown. We did not measure factors such as muscle activity during the head correction exercise, and the mechanism of hypercorrection in the NSNP group is uncertain. In addition, the relationship between neck pain and hypercorrection in the corrected head posture is unknown. Further studies may clarify the cause of hypercorrection in patients with NSNP by measuring muscle activity during head correction, and a cohort study of patients with NSNP to establish a causal relationship between hypercorrection, dysfunction of the deep neck flexors, and neck pain.

## Conclusions

The head and neck alignment in natural and corrected head postures were compared in patients with NSNP and controls, and the relationship between the degree of change and the function of the deep cervical flexors was analyzed. Hypercorrection in the corrected head posture was shown among patients with NSNP; this may be correlated with dysfunction of the deep cervical flexors, and clinicians should consider this when treating these patients.

## Data Availability

All data produced in the present study are available upon reasonable request to the authors

## List of Abbreviations

BMI: body mass index
CCFT: craniocervical flexion test
CRA: cranial rotation angle
CVA: cranial vertical angle
FHP: forward head posture
ICC: intraclass correlation coefficient
NRS: numerical rating scale
NSNP: nonspecific neck pain
PI: performance index

